# High frequency of Exon 20 S768I *EGFR* mutation detected in malignant pleural effusions: a poor prognosticator of NSCLC?

**DOI:** 10.1101/2019.12.08.19014167

**Authors:** George D’Souza, Chirag Dhar, Vishal Kyalanoor, Lokendra Yadav, Mugdha Sharmra, Mohammad Nawaz S, Sweta Srivastava

**Author notes:** Corresponding authors: Dr. Sweta Srivastava-, Dr. George D’Souza- and Dr. Chirag Dhar- /. Equal contribution of authors (George D’Souza, Chirag Dhar).

## Abstract

Lung cancer is the cause of a fourth of all cancer-related deaths. About a third of all lung adenocarcinoma tumours harbour mutations on exons 18, 19, 20 and 21 of the epidermal growth factor receptor (*EGFR*) gene. Detection of these mutations allows for targeted therapies in the form of EGFR Tyrosine kinase inhibitors. In our study, we utilized malignant pleural effusions (MPEs) as “liquid biopsies” to detect *EGFR* mutations when tissue biopsies were unavailable. We showed that a direct sequencing approach was likely to miss SNVs in MPEs. We then optimized an *EGFR* mutant-specific quantitative polymerase chain reaction-based assay and piloted it on n=10 pleural effusion samples (1 non-malignant pleural effusion as a negative control). 5/9 (55.55%) samples harboured EGFR mutations with 2/9 (22.22%) being exon 19 deletions and 3/9 (33.33%) had the S768I exon 20 mutation. The frequency of the S768I SNV in our study was significantly higher than that observed in other studies (∼0.3%). Utilizing publicly available cBioPortal data, we report that patients with the S768I SNV had a shorter median survival time, progression-free survival time and lower tumor mutation count compared to patients with other *EGFR* mutations. These data suggest that this point mutation predicts poor prognosis as a result of aggressive disease, though studies in larger cohorts are necessary to confirm these findings. The high frequency of S768I mutations seen in our study also suggests that cancer cells harbouring these mutations may be superior in their ability to migrate, home or reside in pleural fluid.

## Introduction

Lung cancer is a major contributor to death due to cancer. Amongst the various types of lung cancer, non-small cell lung cancer (NSCLC) accounts for approximately 80% of all cases(1). A significant rise has been seen in the time trends of lung cancer in the Indian cities of Delhi, Chennai and Bangalore. Lung cancer is the cause of 6.9 percent of all new cancer cases and 9.3 percent of all cancer related deaths in both men and women in India (2). Mutations seen on Exons 18, 19, 20 and 21 of the Epidermal growth factor receptor (*EGFR*) gene are frequently observed in these cases and have been shown to be present in nearly a third of all lung adenocarcinoma cases (3)(4)(5). *EGFR* is a family member of receptor tyrosine kinases that play a central role in cellular signalling promoting cell growth and proliferation. Some mutations in this protein strongly predict the efficacy of *EGFR* tyrosine kinase inhibitors used in the treatment of these cases. There are reportedly more than 32 different mutations that have been detected in this gene distributed across exons 18, 19, 20, and 21 (6)(7). More than 20 different in-frame deletions on exon 19 have been reported accounting for nearly half of the cases of *EGFR* mutant lung adenocarcinoma. Importantly, tumours harbouring these deletions are found to be sensitive to EGFR TKIs such as erlotinib and gefitinib (8). This targeted therapy has been found to increase clinical outcomes in these patients (9). Response rates of more than 70 percent have been observed in patients on EGFR targeted therapy (10). Patients with EGFR mutant positive advanced NSCLC treated with Erlotinib, an EGFR Tyrosine kinase inhibitor (TKI) have shown better response rates and Progression-free survival (PFS) as compared to those on first line chemotherapeutic agents (11)(12)(13)(14).

Diagnostic tests for the detection of these EGFR mutations are now part of the routine management of lung adenocarcinoma(15). In spite of the invasiveness and morbidity associated with lung biopsies(16), the primary tumour is preferred for detection of mutations(17). Most often, an amplification-refractory mutation system (ARMS) is used for the detection of these mutations from the biopsy. The utility of malignant pleural effusion (MPE) in the detection of EGFR mutations has been well established over the years in the field of lung cancer diagnosis(18). In this study, we optimized a method to detect EGFR mutations in MPEs at our tertiary care centre. Notably, the single nucleotide variant (SNV) S768I was found in 3 out of the 9 pleural effusions (33.33%). We then analysed the prevalence and clinical features of patients harboring this SNV by utilizing cBioPortal (www.cbioportal.org).

## Results

### Dilutions of mutant in wildtype EGFR DNA predict poor sensitivity of direct sequencing-based detection of SNVs in MPEs

First, a direct sequencing-based approach was optimized to detect EGFR mutations from MPEs. A common exon 19 deletion was detected using this method (figure 1 a and b). We then asked if this method would be suitable to pick up mutant alleles in the backdrop of wildtype inflammatory cells present in MPEs. To answer this question, we serially diluted DNA with the exon 19 deletion with wildtype DNA and subjected the mixture to direct sequencing. While signatures of the large 18bp deletion were visible even in 1:100 dilutions, the results suggested poor base-call confidence (figure 1 c). This result suggested that the direct sequencing method was unlikely to pick up SNVs.

**Figure 1:**
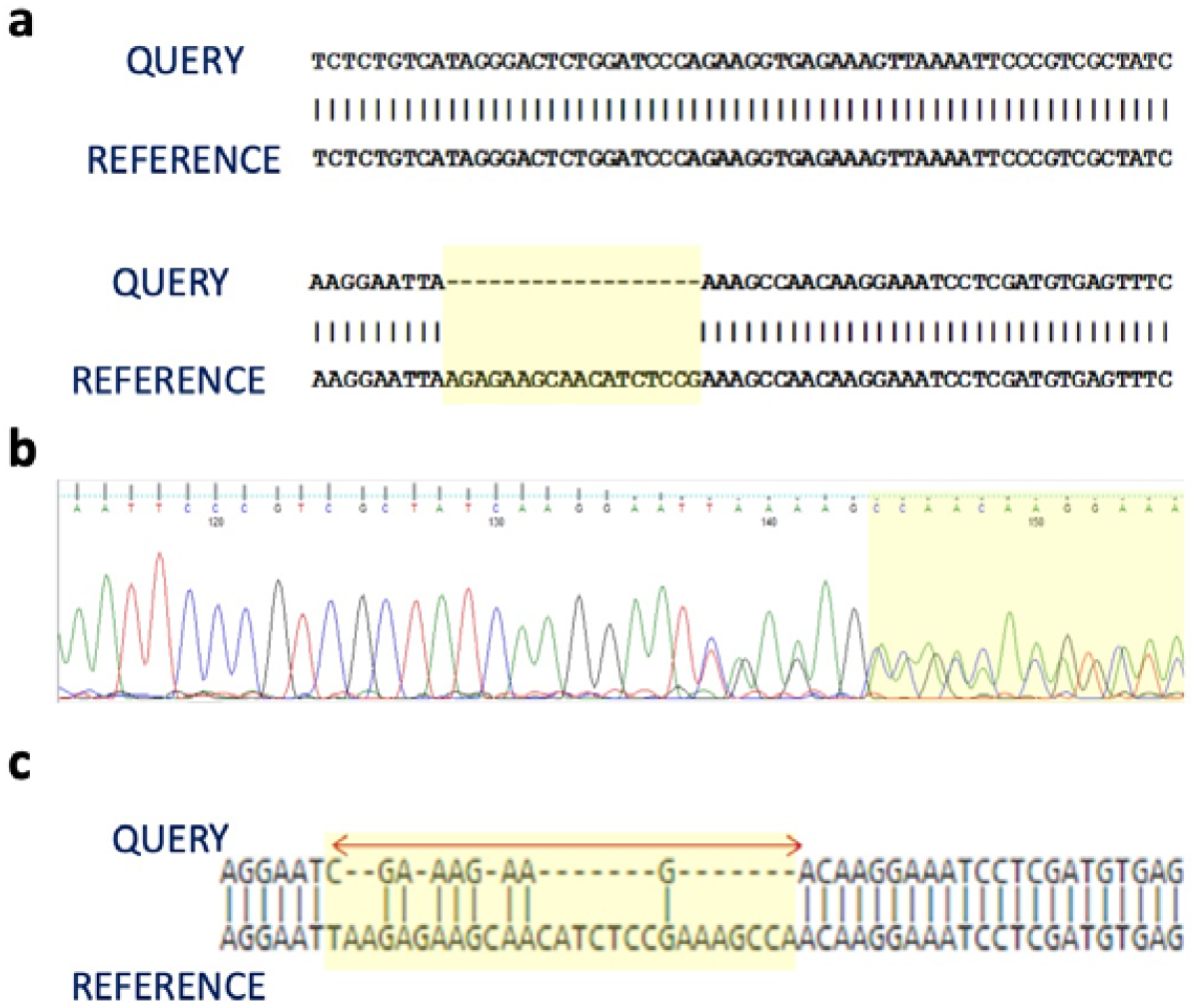
Poor base-call confidence in diluted DNA samples. **a) Common exon 19 deletion detected.** Alignment of MPE sequence with reference sequence shows an 18bp deletion (highlighted in the yellow box). **b) Chromatogram view of the same mutation** (highlighted in the yellow box) **c) Alignment of diluted DNA sample**. Poor base-call confidence in sequence of 1:100 dilution of mutant DNA in wildtype DNA (healthy laboratory volunteer).

### High frequency of S768I exon 20 mutation detected by mutant-specific quantitative PCR of MPE

Subsequently, an established *EGFR* mutation real-time PCR kit was used to identify EGFR mutations in malignant pleural effusions. While this kit is optimized to detect mutations in genomic DNA obtained from primary tumors, we attempted to utilize MPEs instead. We demonstrated for the first time that this kit could detect EGFR mutations from MPEs. Our results showed that 5/9 (55.55%) of pleural effusions probed had EGFR mutations. Of these, 2/9 (22.22%) were the exon 19 deletion E746_S750del18 and 3/9 (33.33%) were the exon 20 S768I mutations (Figure 2 a and b). The amplification plots and ΔCt values are provided in supplementary figures 1-7 and supplementary tables 1 and 2. The occurrence of this SNV and clinical features of these patients was then studied using publicly available data on cBioPortal. This SNV has been detected only 17 times in the nearly 5836 (0.3%) non-small cell lung cancer samples surveyed for *EGFR* mutations (Figure 2 c). These detections include duplicates and the frequency at the patient level is 0.18% (10/5490 patients). This SNV is mapped onto the tyrosine-kinase domain of *EGFR* and is a known cancer hotspot (statistically significantly recurrent mutations identified from large scale cancer genomic studies) (Figure 2 d). Patients with this mutation are suitable candidates for the FDA-approved targeted therapy Afatanib.

**Figure 2:**
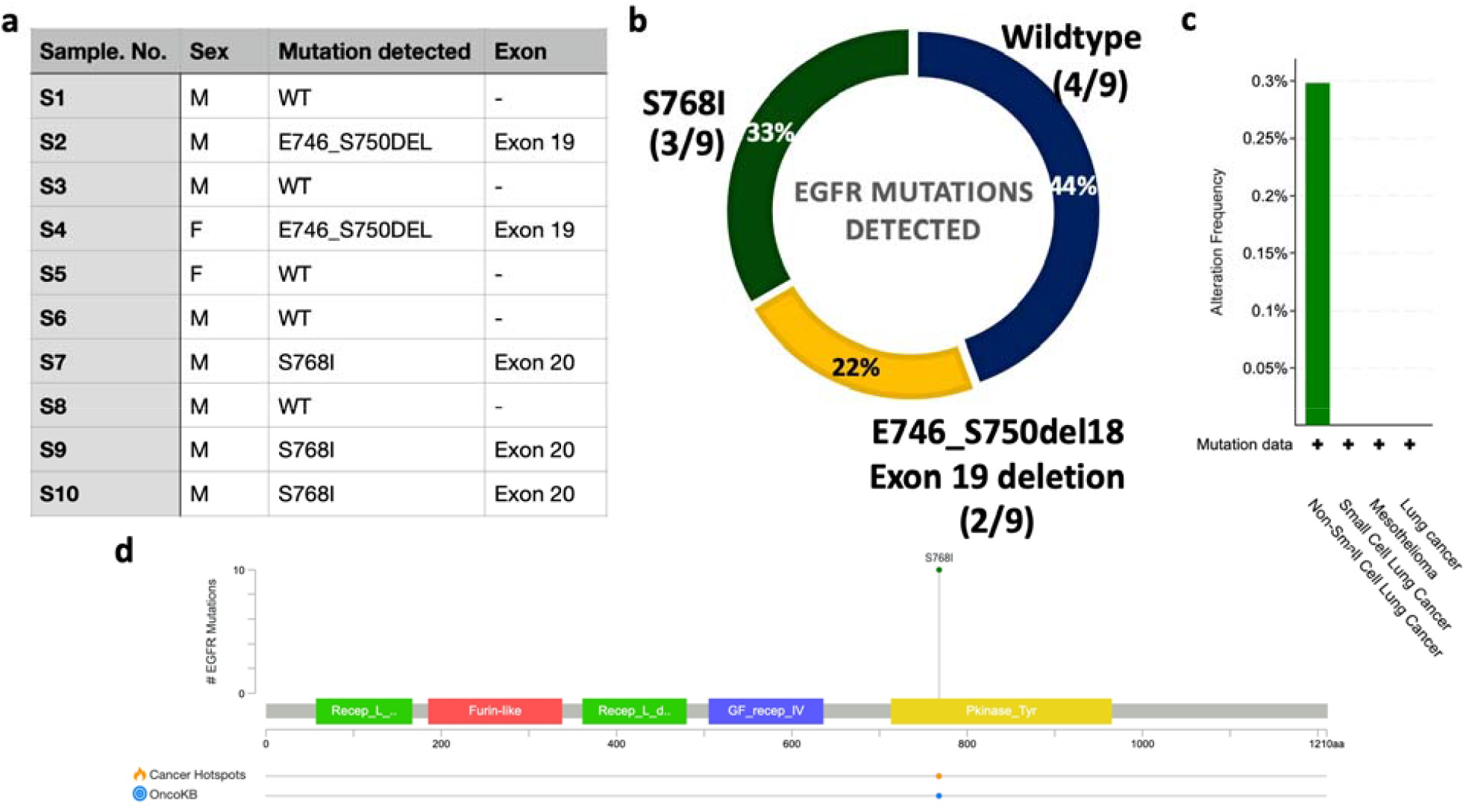
High frequency of S768I detected from MPEs. **a) Table depicting gender, mutation detected and exon of occurrence.** M= Male, F= Female, WT= wildtype *EGFR*, S5 was a non-malignant pleural fluid sample that served as an internal assay negative control **b) Doughnut chart representing the frequency of various mutations detected in this study**. Percentages are embedded in the slices and number of cases observed are indicated in parenthesis beneath the genotype. **c) Frequency of S768I in other studies**. cBioPortal data shows that this SNV has been detected only 17 times at a frequency of 0.4%. **d) Mapping of S768I**. S768I maps to the tyrosine kinase domain of *EGFR* and is a known cancer hotspot.

### Patients having the S768I mutation have a shorter median survival time and a shorter progression-free survival time compared to patients having other *EGFR* mutations

From publicly available cBioPortal data, patients with the S768I mutation have a median survival of 6.20 months compared to 38.40 months patients with other *EGFR* mutations (figure 3 a, b and c). Additionally, patients with the SNV had a shorter progression-free survival times (8.55 months for patients with S768I vs 44.02 months for patients with other *EGFR* mutations, figures 4 a, b and c). These survival statistics (limited by number of patients) suggest that patients with this SNV present with an advanced form of NSCLC.

**Figure 3:**
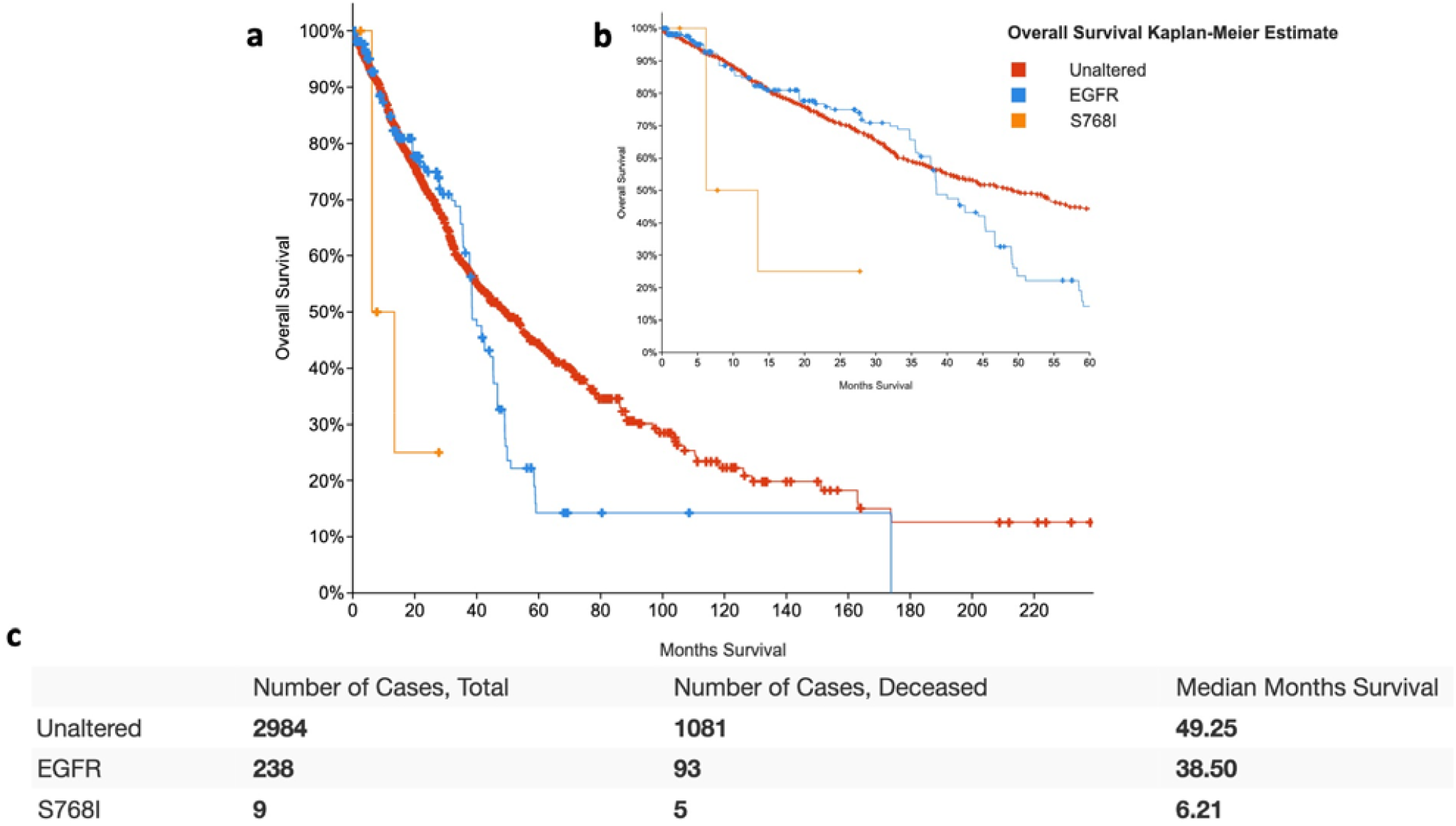
Patients having the S768I mutation have a shorter median survival time compared to patients having other *EGFR* mutations. **a) Kaplan-Meir (KM) survival curve b) KM curve for first 60 months inset c) Median survival in months** suggest shorter median survival in patients with S768I. Unaltered refers to NSCLC patients without *EGFR* mutations, EGFR to patients with *EGFR* mutations except S768I and S768I refers to patients with this SNV.

**Figure 4:**
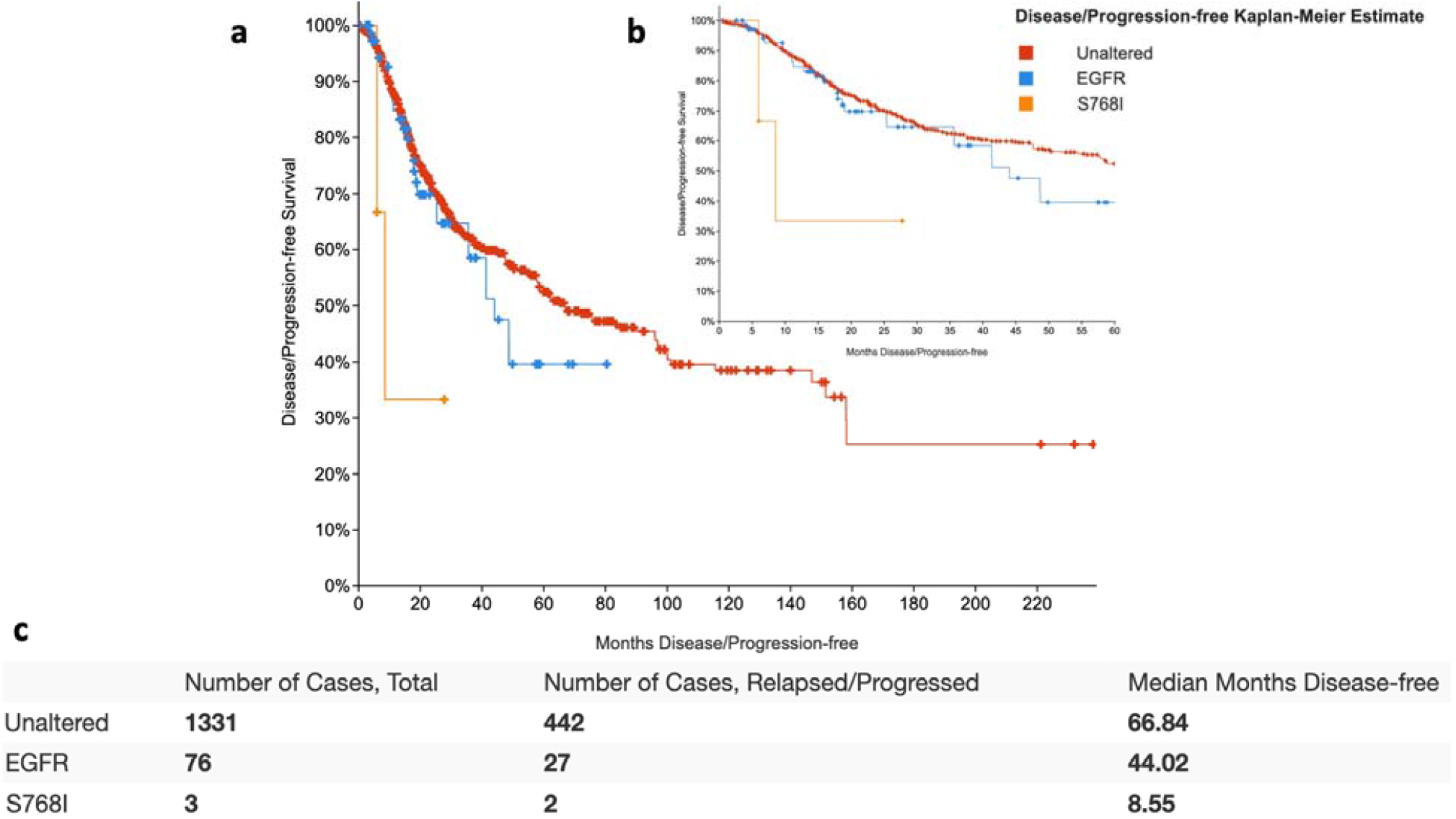
Patients having the S768I mutation have a shorter progression-free survival time compared to patients having other *EGFR* mutations. **a) Kaplan-Meir (KM) survival curve b) KM curve for first 60 months inset c) Median survival in months** suggest shorter median time to relapse in patients with S768I. Unaltered refers to NSCLC patients without *EGFR* mutations, EGFR to patients with *EGFR* mutations except S768I and S768I refers to patients with this SNV.

### Presence of the S768I SNV was not associated with a significant difference in stage at diagnosis, lymph node involvement, size of tumor and metastasis

Given the poorer survival in the S768I group, we compared the stage at diagnosis, lymph nodal involvement, size of tumor and metastasis between the groups. There was no difference in the stage at diagnosis between patients with or without the S768I mutation (figure 5 a). There also was no discernible difference in lymph nodal involvement (figure 5 b) or tumor size (figure 5 c) between the two groups possibly due to the small sample size in the S768I group. We then queried the number metastasis samples obtained from these patients as a proxy for frequency of metastasis. Again, there was no difference in the frequency between the groups (figure 5 d). It is likely that a larger meta-analysis of this SNV may reveal discernible differences in these factors but in a limited number of cases with available data this was not the case.

**Figure 5:**
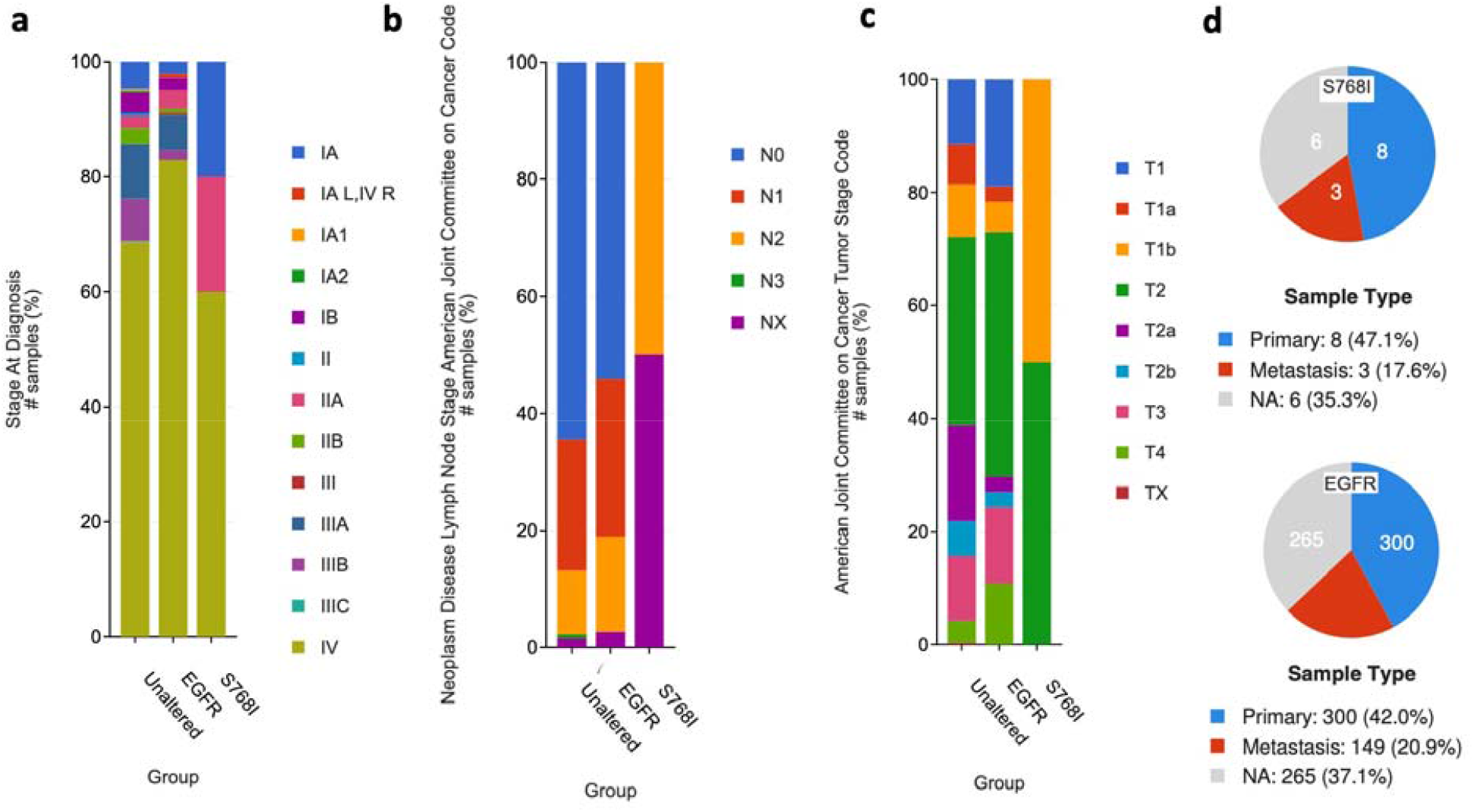
No observable differences in stage at diagnosis, lymph nodal involvement, tumor stage code or metastasis in tumors with S768I. Differences between groups by a) Stage at diagnosis b) Lymph nodal involvement c) Tumor stage code and d) metastatic lesions sampled as a proxy for metastatic frequency. Unaltered refers to NSCLC patients without *EGFR* mutations, EGFR to patients with *EGFR* mutations except S768I and S768I refers to patients with this SNV.

### Tumors with the S768I SNV have a lower mutation burden but co-occur with mutations on other genes

We then asked if patients with the S768I patients had a lower mutation count compared to patients with other *EGFR* mutations. Indeed, the number of mutations was lower in the S768I group (figure 6 a) suggesting a more deleterious effect of this SNV. These data when taken together with the survival data point towards a likely pathogenic role of S768I in advanced NSCLC. Further analysis revealed association of the S768I SNV with other mutations. This SNV was as likely to co-occur with TP53 mutations as tumors with no EGFR mutations or ones with other EGFR mutations. Yet, S768I co-occurred with mutations on other genes such as MUC17, BMPR2, DOCK3, TPTE, ATRX and FSHR (figure 6 b, c and d). The clinical significance or mechanistic importance of these co-occurring mutations is unknown.

**Figure 6:**
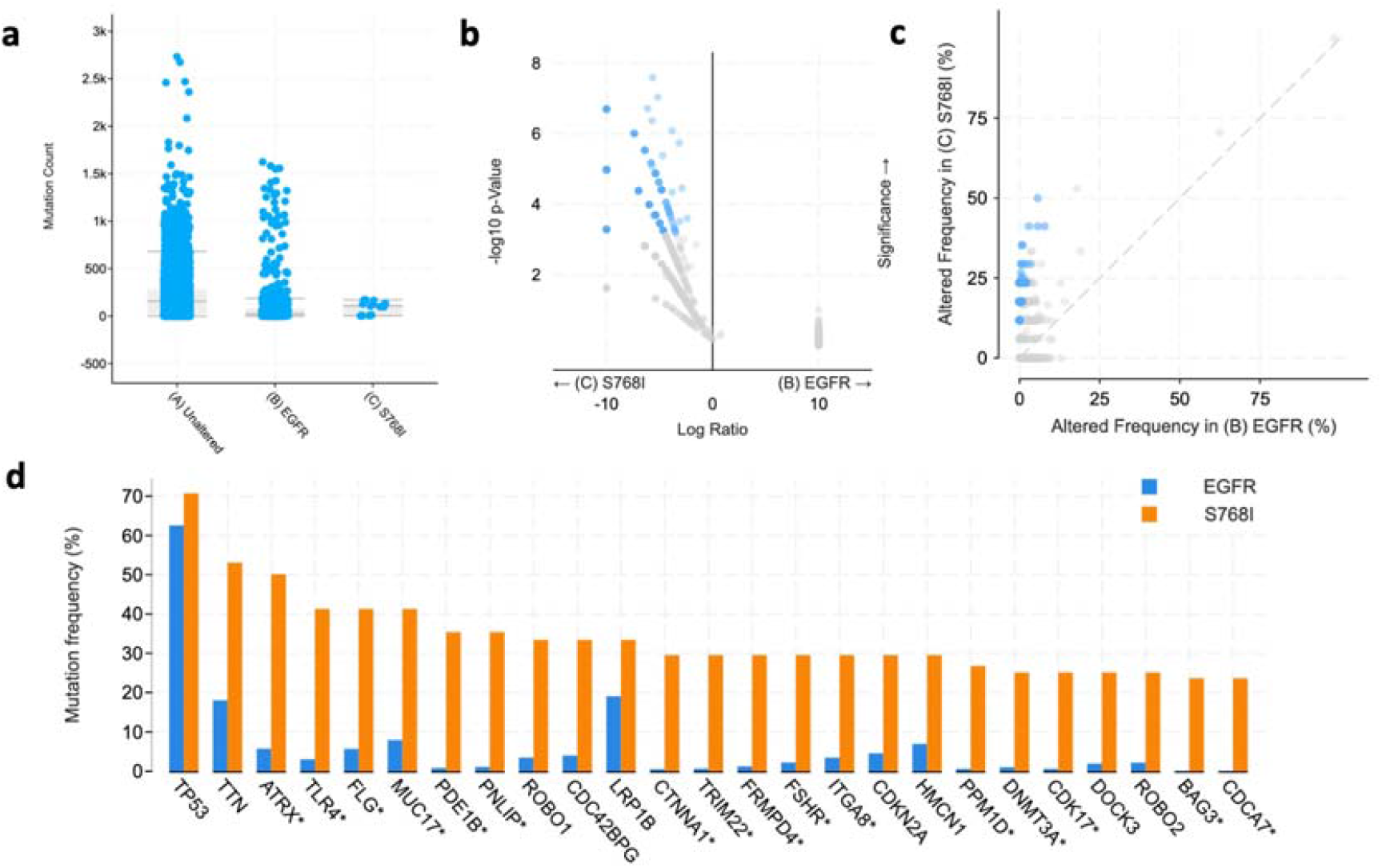
Lower tumor mutation in patients with S768I and co-occurrence with other mutations. **a) Mutation load in tumors with S768I.** The overall mutation load in tumors with the S768I SNV is lower than in tumors with other EGFR mutations or NSCLC samples with other mutations. **b) Graphical representation of genes mutated more frequently in tumors positive for S768I but not in tumors with other EGFR mutations. c) Dot plot depicting frequency of co-occuring mutations in S768I tumors**. Blue dots represent genes with a statistically significant p-value. **d) Bar graphs of 25 most frequently detected mutations (apart from EGFR)**. TP53 mutation frequency is nearly the same but mutations in genes such as FLG, MUC17, BMPR2, DOCK3, TPTE, ATRX and FSHR mutations appear more frequently in conjunction with the S768I mutation. Asterisked (*) genes represent those with statically significant differences. A list of all 175 genes with statistically significant co-occurrence with S768I are listed in the supplementary information.

## Discussion

In this study, we showed that a direct sequencing approach may miss *EGFR* SNVs in MPEs and therefore utilized a Taqman-based mutant-specific quantitative PCR assay. We optimized this assay and detected both large deletions and SNVs in *EGFR*. Notably, a high frequency of the S768I mutation occurring in the absence of other *EGFR* mutations was seen. We then compared this frequency to publicly available data on cBioPortal ® and observed a significant difference in frequency (33.33% in our study vs 0.4% in reported literature). To further understand the role of this SNV, a cBioPortal ® search and comparison strategy was devised to compare the clinical features of patients with the SNV, patients with other *EGFR* mutations and patients with no mutaions in *EGFR*. Patients with the S768I had an 84% reduction in estimated median survival time (and progression-free survival time) compared to those with other *EGFR* mutations. Tumors bearing the S768I mutation also had a lower mutational than the other *EGFR* mutations group. These data when taken together suggests that patients with the S768I mutation have poorer clinical outcomes and this mutation is likely to promote an aggressive tumor phenotype.

Very little is known about the implications of having the S768I mutation in the absence of other *EGFR* mutations. OncoKB suggests the use of afatinib as the FDA-approved therapy for patients with this SNV. One other study shown that S768I conferred reduced sensitivity to gefitinib *in vitro* as compared to other mutations (19)(20). A study by Leventakos and group reported that the S7681 is rare and is usually found in combination with sensitizing mutations and they have concluded that the predictive and prognostic role of this mutation is yet to be fully explored (21). In our study we have found that 33.3% of the samples exhibited this mutation in isolation, with an absence of any other mutation. It did not escape our attention that all reports of this SNV in studies discussed here occur in the presence of other EGFR mutations (such as at the G719 or T790 position). Some of the reasons for this mutation being detected in the absence of other *EGFR* mutations in our study are discussed here. Most studies utilize the primary tissue and a sub-population with this isolated mutation may appear later in the pathogenesis of the disease. One may also simply come to the conclusion that our cohort has a larger number of patients with this mutation, but we think otherwise. We hypothesize that S768I is a secondary mutation that develops later in the course of the disease and cells harbouring this mutation are able to migrate to, home in or reside within the pleural cavity more easily that cells without this mutation. One could further this speculation that cells having this mutation have a greater metastatic potential (figure 7) though further studies will be needed to prove these hypotheses. Additionally, *in vitro* and *in vivo* studies will determine if this mutation is likely to respond to EGFR TKIs such as afatinib.

**Figure 7:**
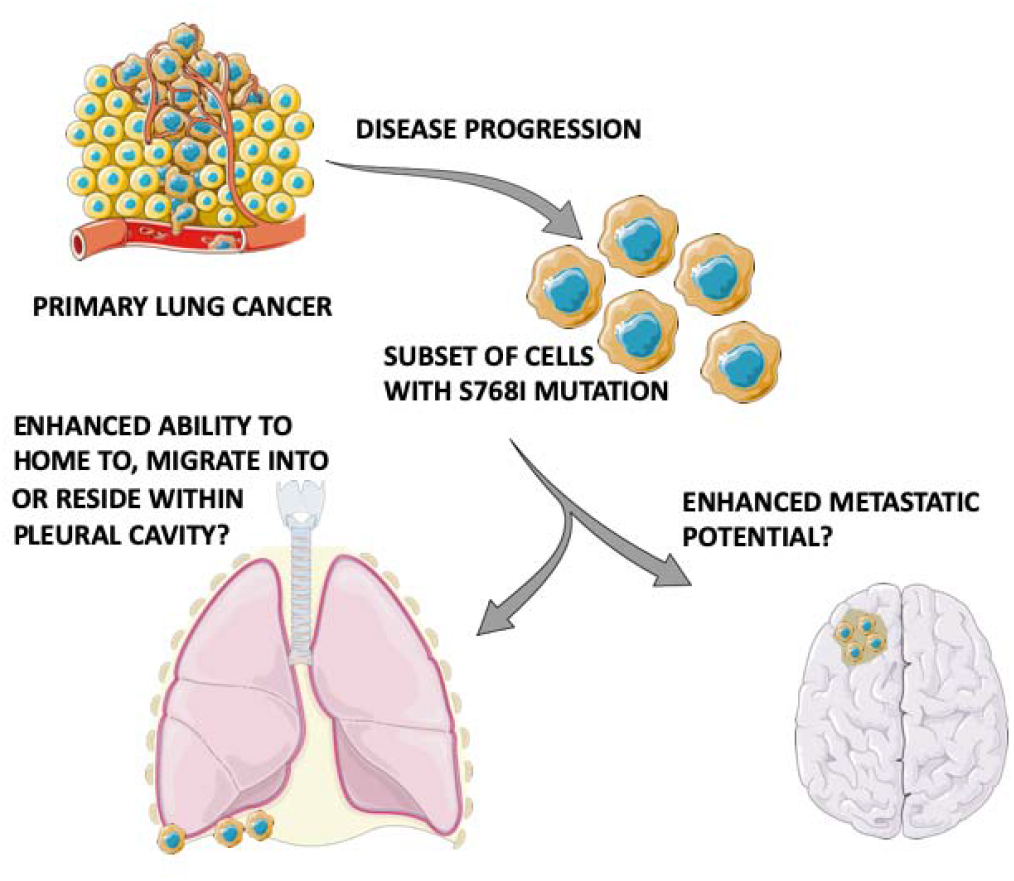
Hypothesized role of the S768I mutation in the pathogenesis of metastasis in lung cancer (Elements of images sourced from http://smart.servier.com/).

The prevalence of somatic mutations in the *EGFR* gene is high (∼ 33%) in lung adenocarcinomas(22)(23). The detection of any *EGFR* mutation and the resultant treatment with targeted therapy with EGFR TKIs (Tyrosine Kinase Inhibitors) aid in sensitizing these tumours. *EGFR* mutation testing on tissue biopsies is the gold standard for diagnosis. However, biopsy procedures may result in uncontrolled bleeding and morbidity(24). Utility of pleural effusion fluid for mutation testing along with other secondary specimens (pericardial effusion, fine needle aspirates, CSF, cfDNA, etc) have been used as alternate strategies for reliable testing (25). In our study we first used PCR coupled to direct sequencing to detect mutation in pleural effusion derived malignant cells. We were able to detect a common 18bp deletion mutation using this method. One of the biggest drawbacks of direct sequencing versus allele specific detection lies in its limit of detection. Pleural fluid consists of a heterogeneous pool of stromal and inflammatory cells apart from the metastatic cells. Since the non-tumor cells harbour wildtype genomic sequence, the signal to noise ratio in electropherogram is lower for mutated sequence. The mutation sequence identification may be compromised in the event of suboptimal malignant cells numbers. We showed that at dilution of 1:100 of a mutated DNA with wildtype genomic DNA signatures of the mutation are visible but the wildtype sequence bases provide conflicting results in certain positions of the gene. These inadequate results are more likely to be seen in single base pair mutations. We then utilized an *EGFR* mutation detection kit. This diagnostic kit is designed to test for *EGFR* mutation on FFPE sections but we were able to optimize its use on MPEs. To the best of our knowledge, this is the first report of MPE *EGFR* mutation testing in India utilizing the Taqman technology platform. Jong Sik Lee and colleagues have used PNA clamping technology to detect EGFR mutations in the supernatant derived from the pleural effusion where in the objective was to detect mutations using cell free DNA (26). Jie Lin et al have shown that EGFR mutations can be detected in the supernatant, cell pellets of the pleural effusion using High Resolution Melting (HRM) analysis and Sanger sequencing (27). When implemented on a small pilot scale study (n=9), we were successful in detecting EGFR mutations in 5/9 (55%) subjects. The mutations that were detected in these samples were either S768I or E746_S750del.

The need to develop minimally invasive diagnostic tests in the management of cancers is gaining momentum. However, our study is limited in its impact as it evaluates only 11 patients and further study needs to be undertaken to evaluate and validate this method in a larger cohort. Additionally, the detection of circulating tumour DNA should also be explored as an alternative to repeated biopsies is one of the techniques being studied (28). This detection is being done using highly sensitive techniques such as droplet digital polymerase chain reaction. Resources must be directed towards the development of such minimally/non-invasive techniques, especially if these are inexpensive as well.

### Ethics

Ethical clearance was obtained from the Institutional Ethics Committee of St. John’s National Academy of Health as per IERB guidelines (Study No. 1/2016). Appropriate consents for carrying out experimental genetic tests were obtained.

## Methods

### Pleural fluid processing and DNA isolation

Pleural fluid was tapped from patients with malignant pleural effusions and centrifuged at 3500 rpm for 5 mins at 4°C to collect the cell pellet. The cell pellets were stored in −80°C until further processing. The DNA was extracted from the samples using the QIAamp DNA Mini Kit (Qiagen) as per the manufacturer’s instructions. Samples were analyzed for their quality using BioSpec Nano UV-VIS Spectrophotometer (Shimadzu Corp.) and electrophoresed on an agarose gel to ascertain integrity.

### Direct sequencing

Appropriate primers for exons 18, 19, 20 and 21 of the EGFR gene were designed and obtained based on the reference sequence available on NCBI (Supplementary table 2). Genomic DNA obtained was subjected to a gradient PCR to optimize the annealing temperature and was visualized on a 1.2% agarose gel (Supplementary figure 8). The PCR products were subjected to direct sequencing.

### Real-time PCR based detection of EGFR mutations

DNA samples were subjected to an assay to a detect 10 *EGFR* mutations in a 96-well Real Time PCR format using the TRUPCR® EGFR Kit (3B BlackBio Biotech India Ltd.) as per the manufacturer’s instructions. Real-time PCR data analysis and identification of EGFR mutant samples was done. To the best of our knowledge, this is the first time this kit is being using to detect mutations from MPEs.

### cBioPortal search strategy

A search strategy was used to access publicly available cancer genomic/clinical data on cBioPortal(29)(30) utilizing multiple studies(31)(32)(33)(34)(35)(36)(37)(38)(39)(40)(41)(4)(42) with the gene alias “EGFR: MUT = S768I”, “EGFR” and to access all NSCLC studies. Venn-diagram based sorting was utilized to isolate cases with the S768I mutation and those with other *EGFR* mutations. “Unaltered” refers to patients with mutations other than EGFR, “EGFR” refers to patients with *EGFR* mutations other than S768I and “S768I” refers to patients with the S768I *EGFR* mutation. OncoKB(43) was used to identify any FDA-approved drug for this mutation.

## Data Availability

There is no metadata associated with this study. All relevant data files are included as images or supplementary images.

## Acknowledgements

This work is supported by a grant from the Advanced Research Wing of the Rajiv Gandhi University of Health Sciences, Bangalore. C.D. was awarded a C.R.E.S.T MAS scholarship by CTRI at the School of Medicine, University of California, San Diego. We thank Prof. Sudhir Krishna, Ms. Pranatharthi Annapurna and the sequencing division of NCBS, Bangalore for their support. Some of these data were presented at the National Lung Cancer Conference 2016 in Bhubaneshwar and at the American Society for Cellular Biology-European Molecular Biology Organization joint conference 2018 at San Diego(44)(45)(46). A special mention to Dr. Paul Kalanithi for his battle against EGFR mutant lung cancer. His memoir When Breath becomes Air is humbling and yet, inspiring.

## Author contributions

G.D.S and C.D contributed equally this work which was conceived, planned, supervised and interpreted by G.D.S, C.D and S.S. Funding acquisition was by G.D.S. Experimental assays were performed and interpreted by C.D., V.K., L.Y. and M.S. Clinical sample acquisition was by C.D., M.N.S. and G.D.S. C.D. performed cBioPortal/bioinformatic analysis. Manuscript was written and edited by C.D. and V.K. S.S. edited the manuscript. All authors approved the final manuscript.

## Supplementary Information

**Supplemental figure 1.**
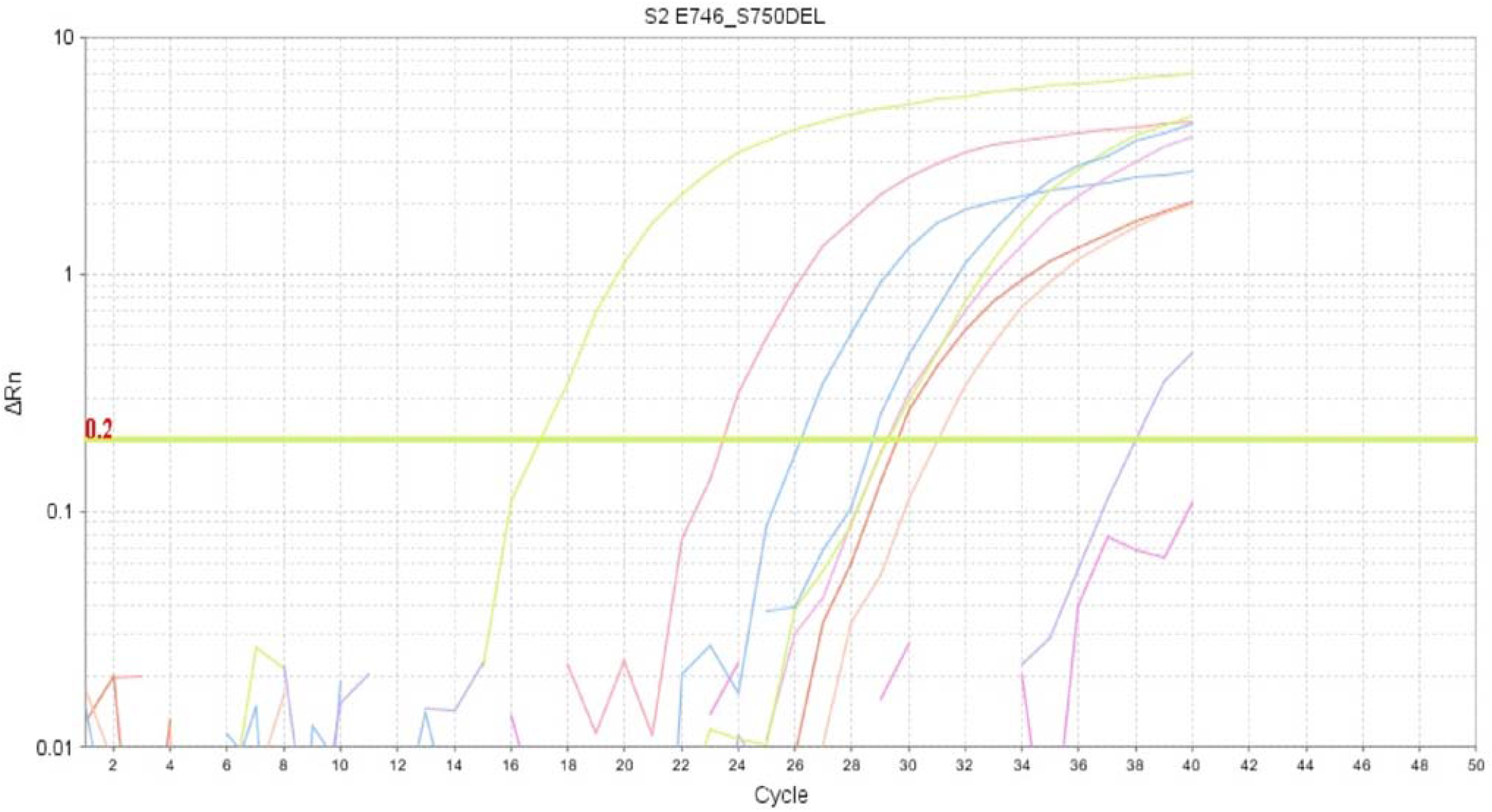
shows the amplification curves for different mutations tested in Sample S2.

**Supplemental figure 2.**
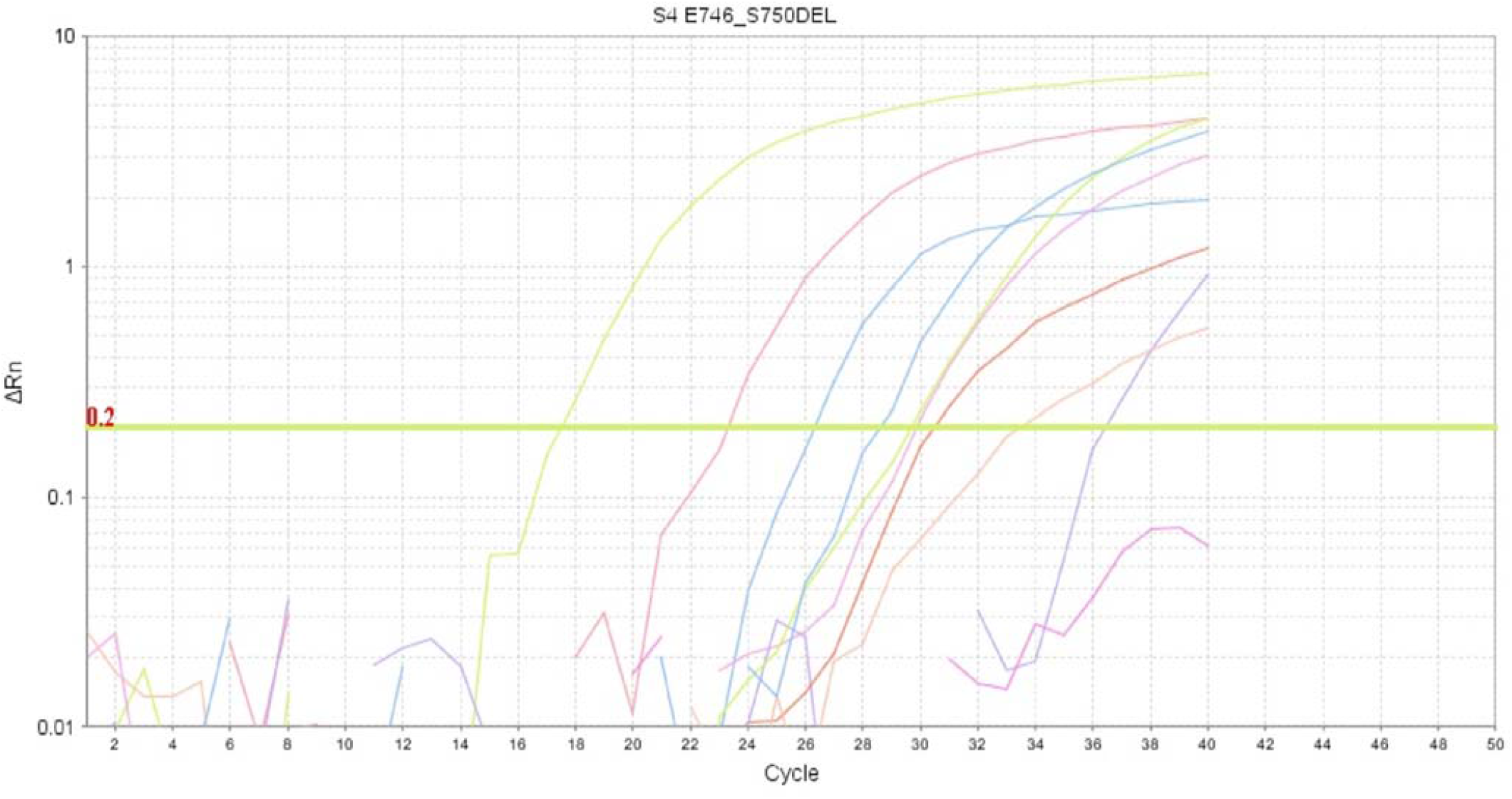
shows the amplification curves for different mutations tested in Sample S4.

**Supplemental figure 3.**
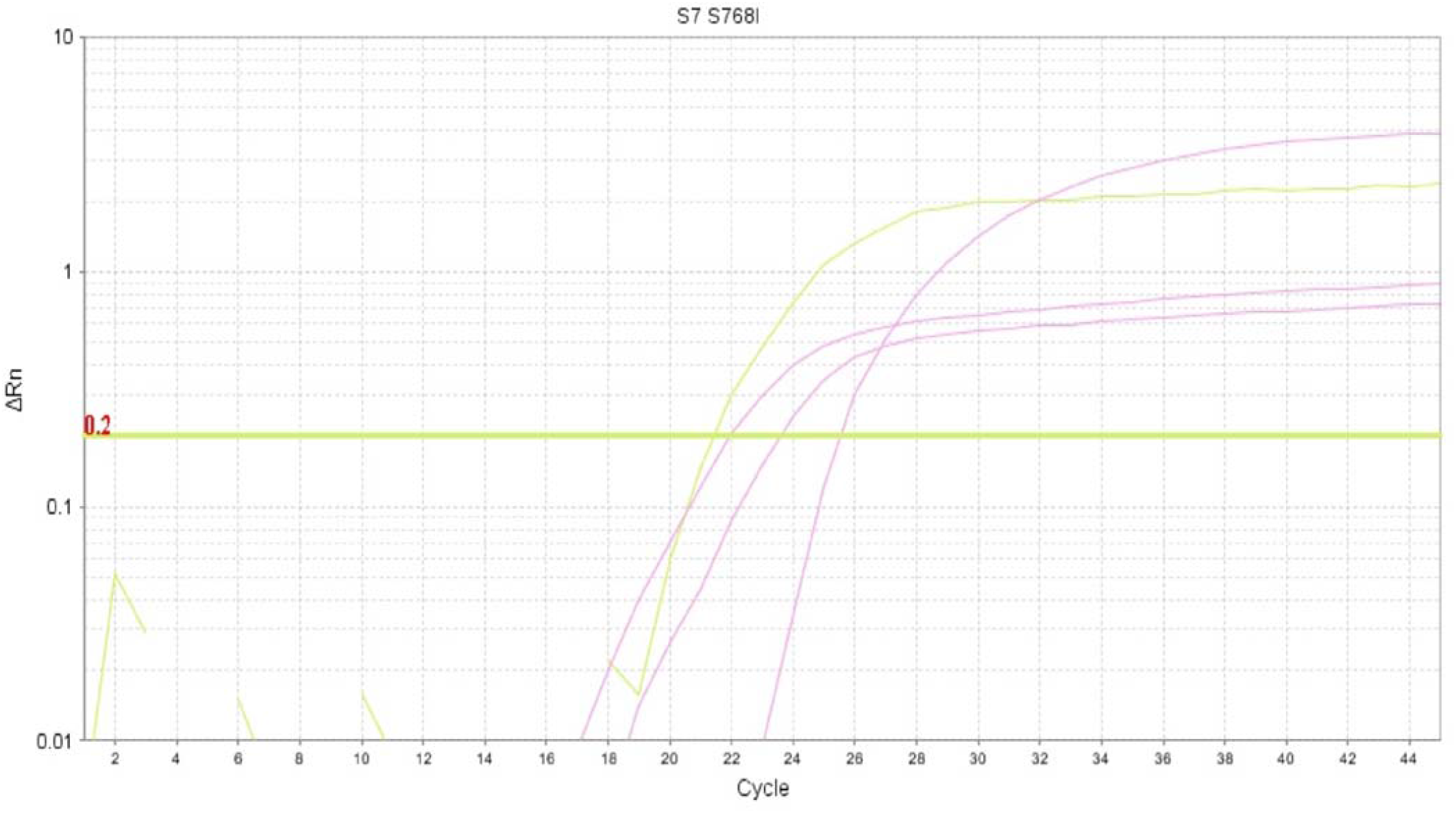
shows the amplification curves for different mutations tested in Sample S7.

**Supplemental figure 5.**
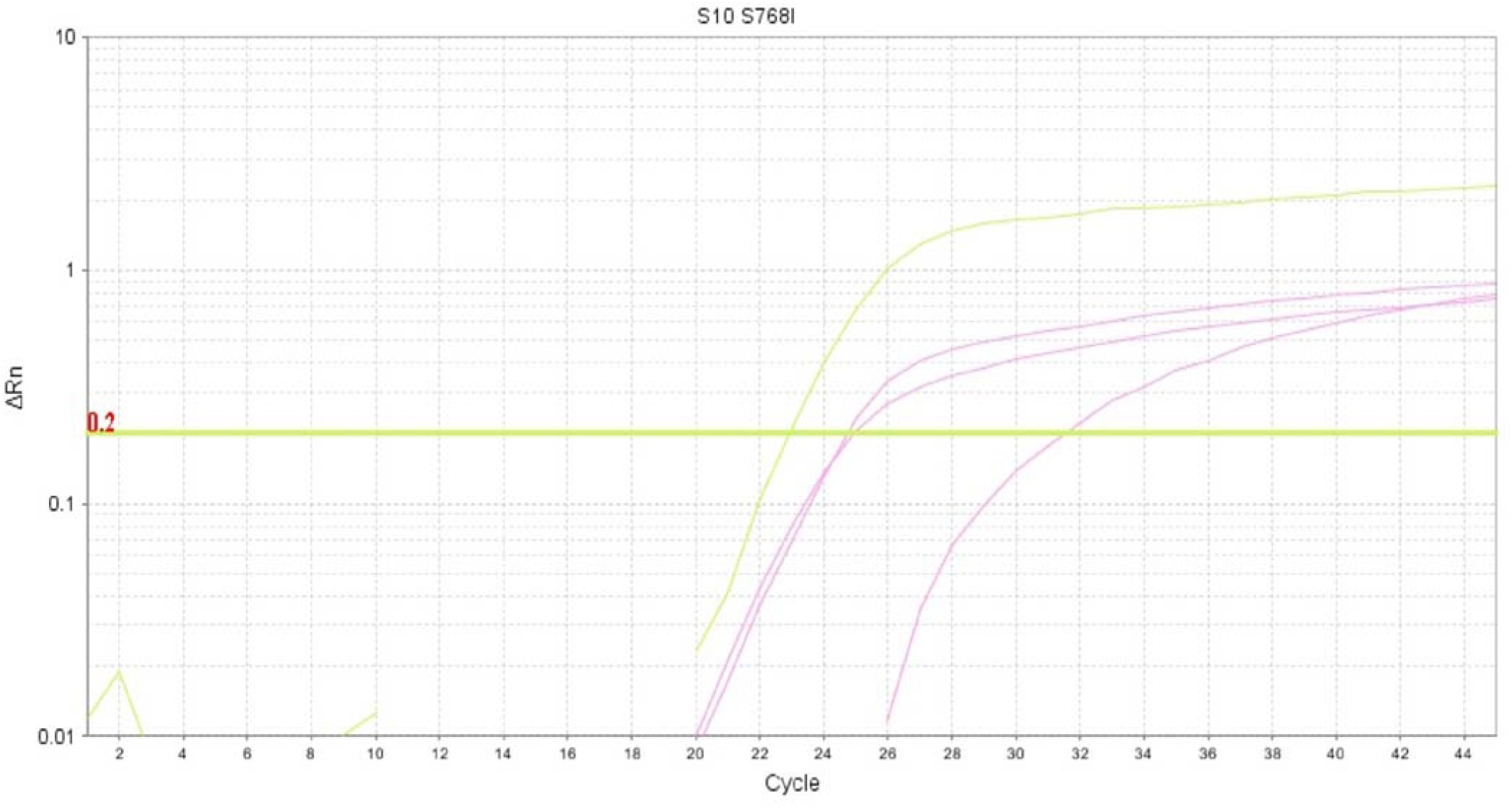
shows the amplification curves for different mutations tested in Sample S9.

**Supplemental figure 6.**
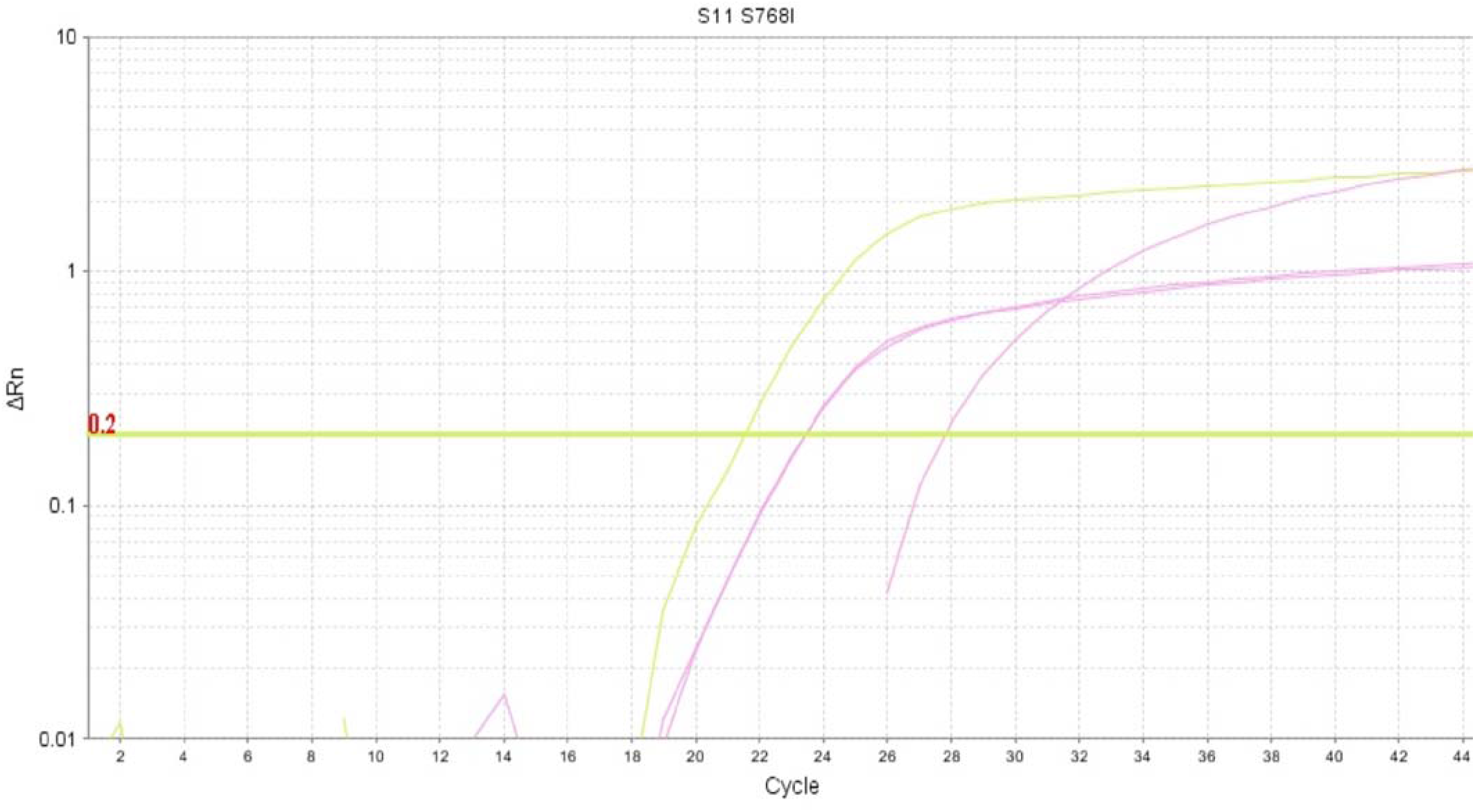
shows the amplification curves for different mutations tested in Sample S10.

**Supplemental figure 7.**
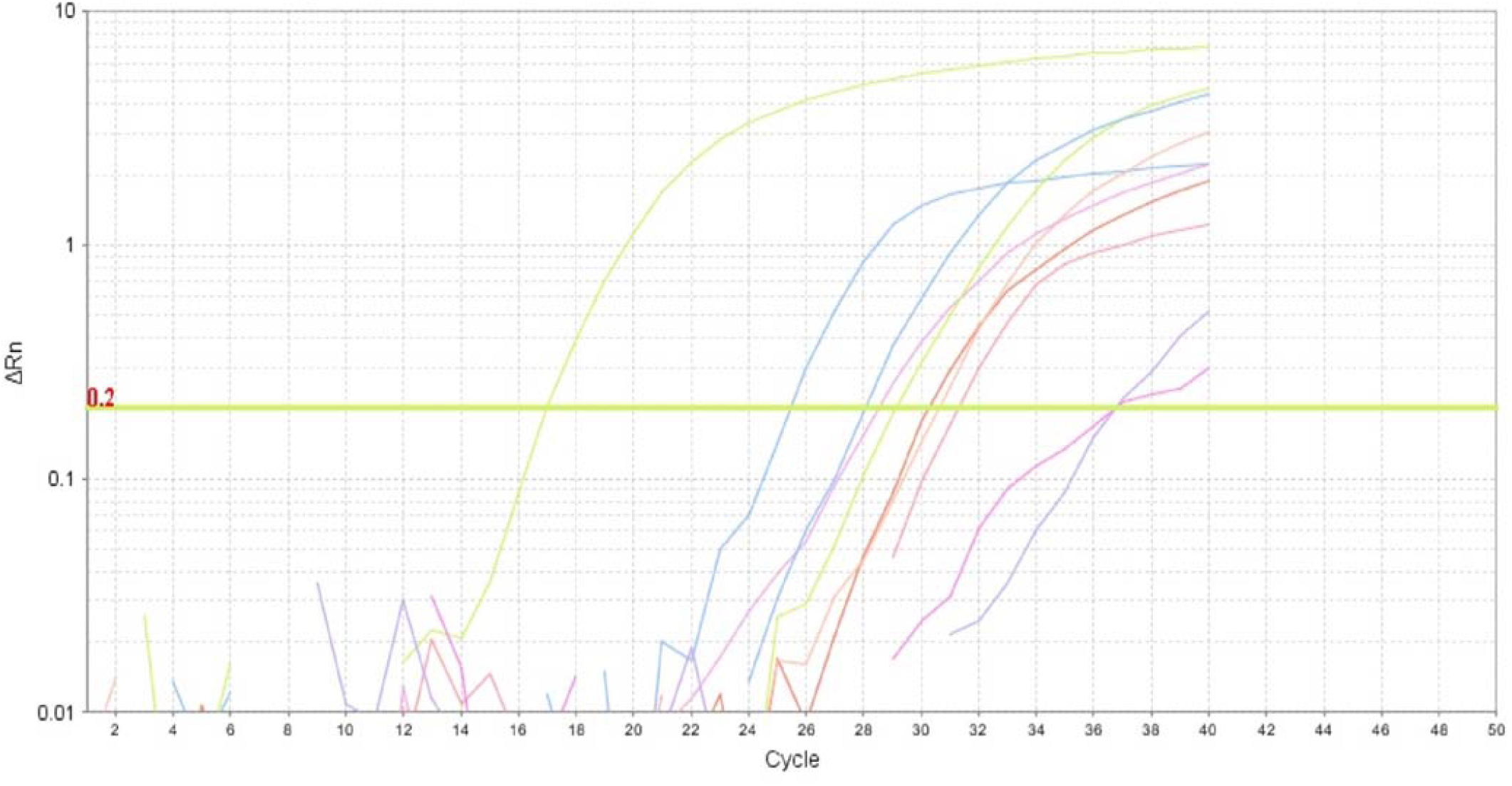
shows the representative amplification curves for different mutations tested and classified as a Wild Type sample or below LOD.

**Supplemental table 1.** shows the ΔCt values of Samples S1-S6.

| Sample No. | ΔCt |  |  |  |  |  |  |  |  |
| --- | --- | --- | --- | --- | --- | --- | --- | --- | --- |
|  | G719X | T790M | S768I | EX20INS | C797S | L858R | L861Q | E746_S750DEL | EX19DEL |
| S1 | 11.261 | 12.919 | 12.393 | 15.518 | 13.704 | 21.527 | 19.990 | 14.589 | 8.697 |
| S2 | 11.595 | 12.284 | 12.148 | 13.915 | 12.470 | 20.835 | ND | 6.327 | 9.159 |
| S3 | 10.997 | 12.154 | 11.452 | 13.572 | 13.223 | 19.834 | 19.788 | 14.197 | 8.416 |
| S4 | 10.981 | 12.106 | 12.276 | 15.961 | 12.879 | 18.905 | ND | 5.597 | 8.640 |
| S5 | 12.438 | 11.107 | 11.385 | 14.670 | 19.424 | ND | ND | ND | 8.957 |
| S6 | 11.314 | 12.079 | 11.776 | 17.363 | 12.901 | ND | ND | 14.173 | 8.642 |

**Supplemental table 2.**
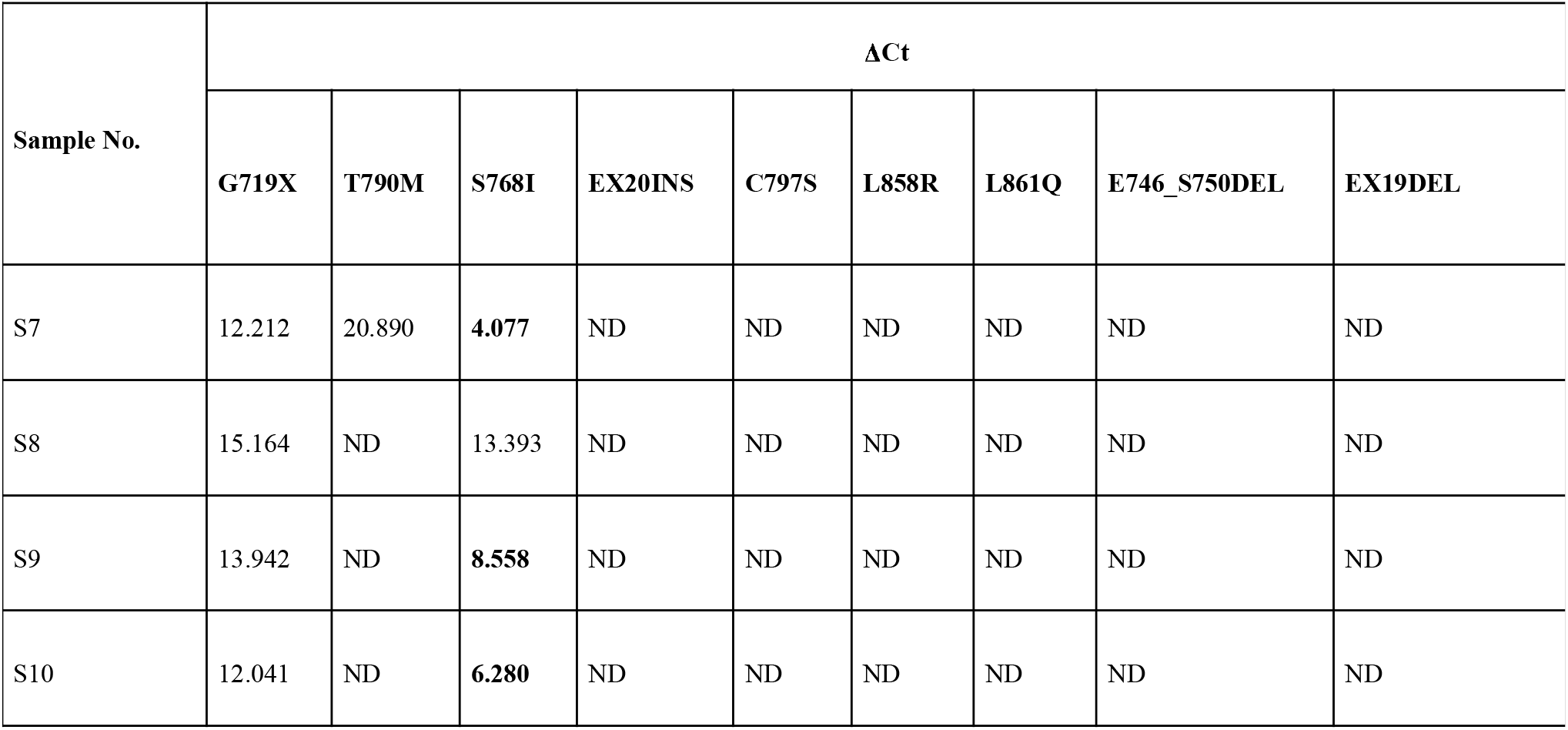
shows the ΔCt values of Samples S7-S11.

**Supplementary table 3.**
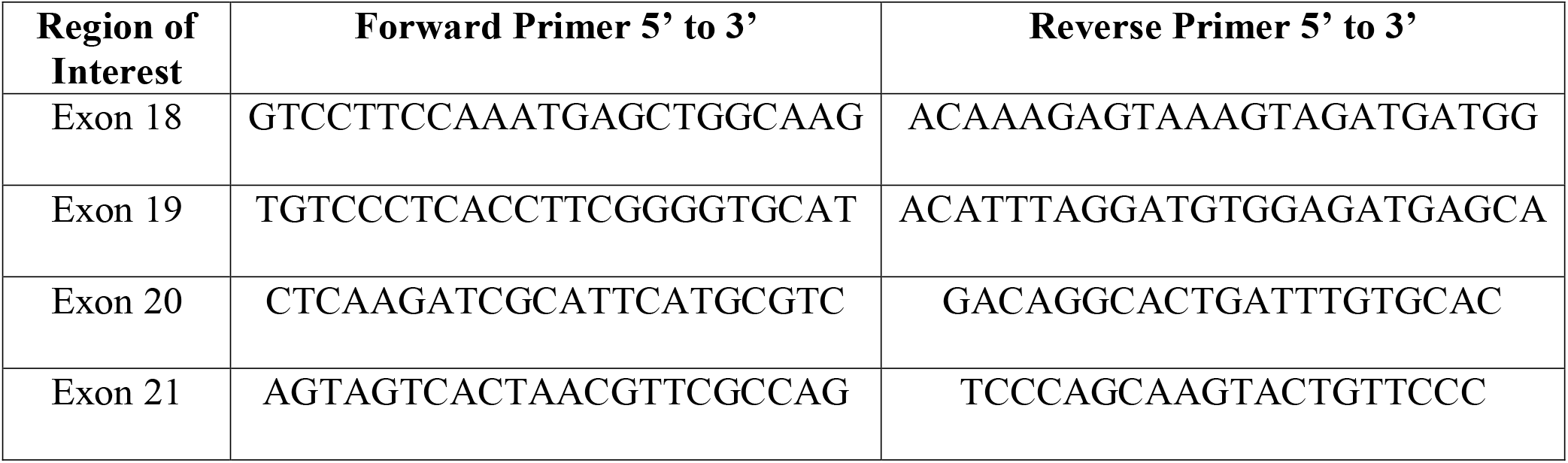
List of newly designed primers

### Optimization of a polymerase chain reaction protocol to amplify EGFR exon 18, 19, 20 and 21 from malignant pleural effusion (MPE)

Initially, we attempted a PCR amplification coupled with a direct sequencing approach to detect EGFR mutations from malignant pleural effusions. Appropriate primers (sequences listed in table 1) were designed and optimized on sample S0. Successful amplification of exons 18-21 by PCR on S0 are shown in figure 1. The primers for exon 18 and 21 had 55 °C as the annealing temperature while exon 19 and 20 were best amplified at an annealing temperature of 57.8 °C.

**Supplementary figure 8:**
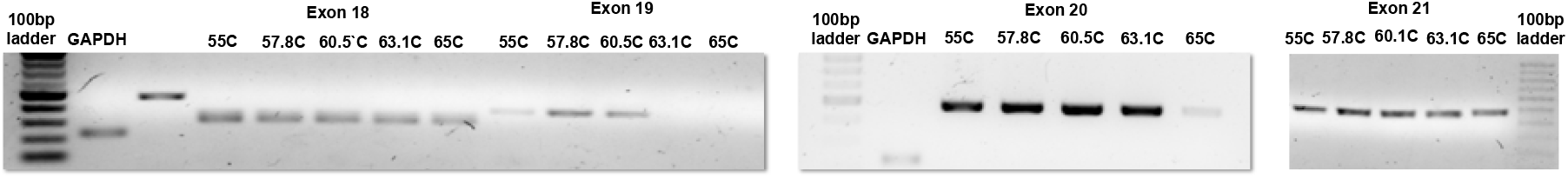
1.2% Agarose gel imaged under ultra-violet light: 100bp ladder, A: GAPDH, B: Exon 20, lanes C-G: Exon 19 at annealing temperatures of 55 C, 57.8 C, 60.5 C, 63.1 C and 65 C lanes H-L: Exon 18 at annealing temperatures of 55 C, 57.8 C, 60.5 C, 63.1 C and 65 C, lanes M-Q: Exon 21 at annealing temperatures of 55 C, 57.8 C, 60.5 C, 63.1 C and 65 C Interestingly, two distinct bands are visible for Exon 19 (lanes C-G)

### List of genes with mutations co-occurring with S768I

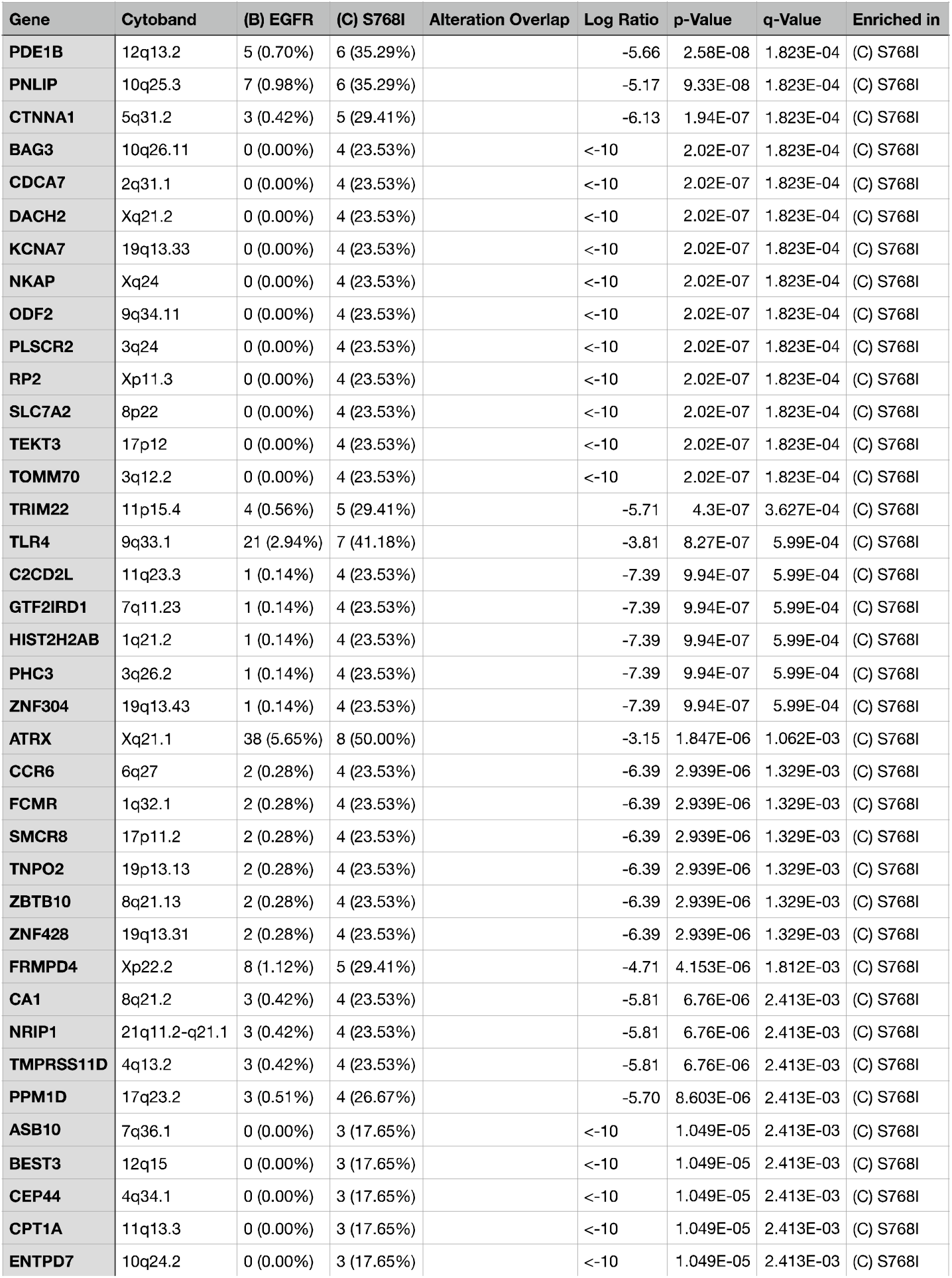
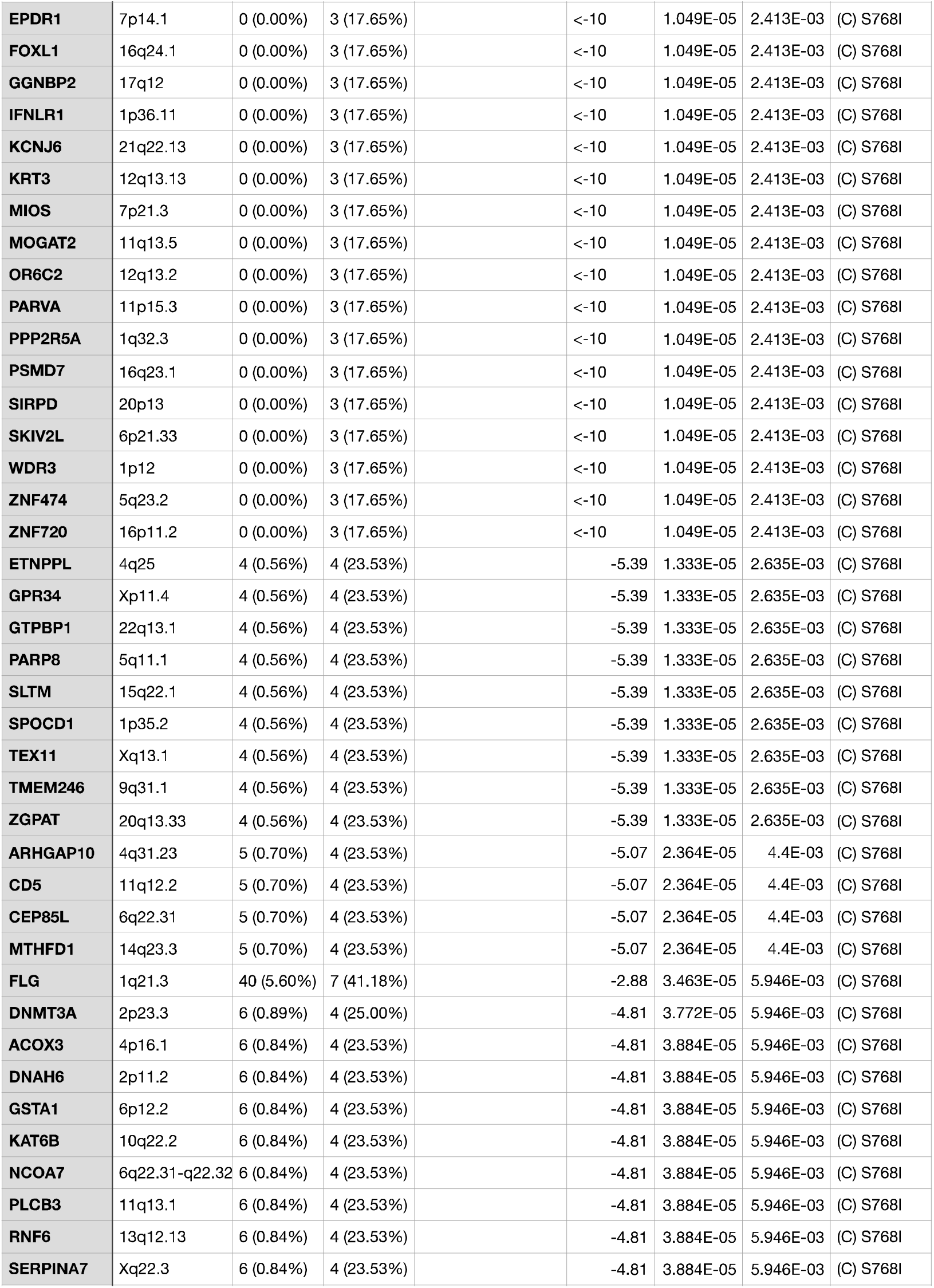
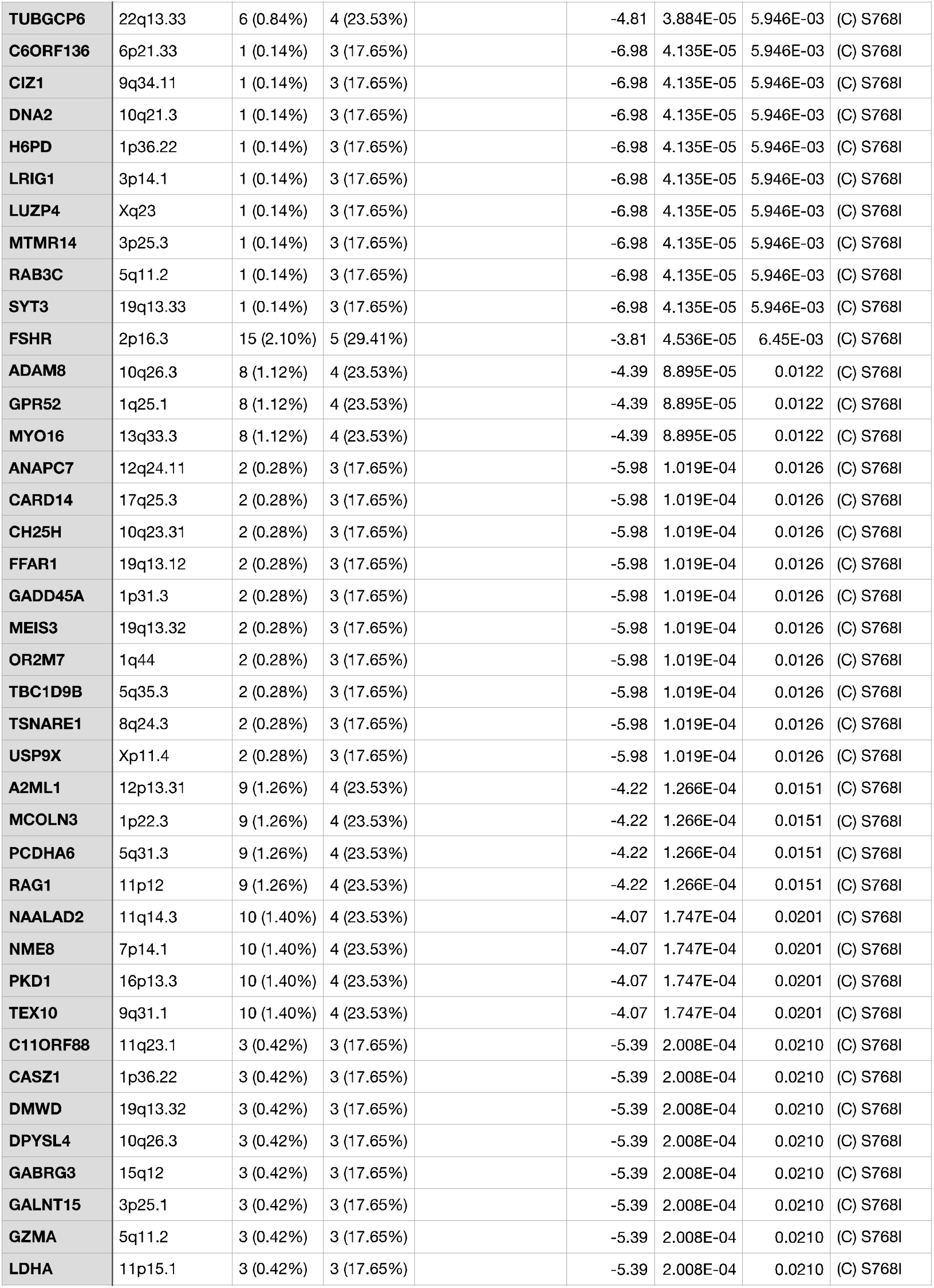
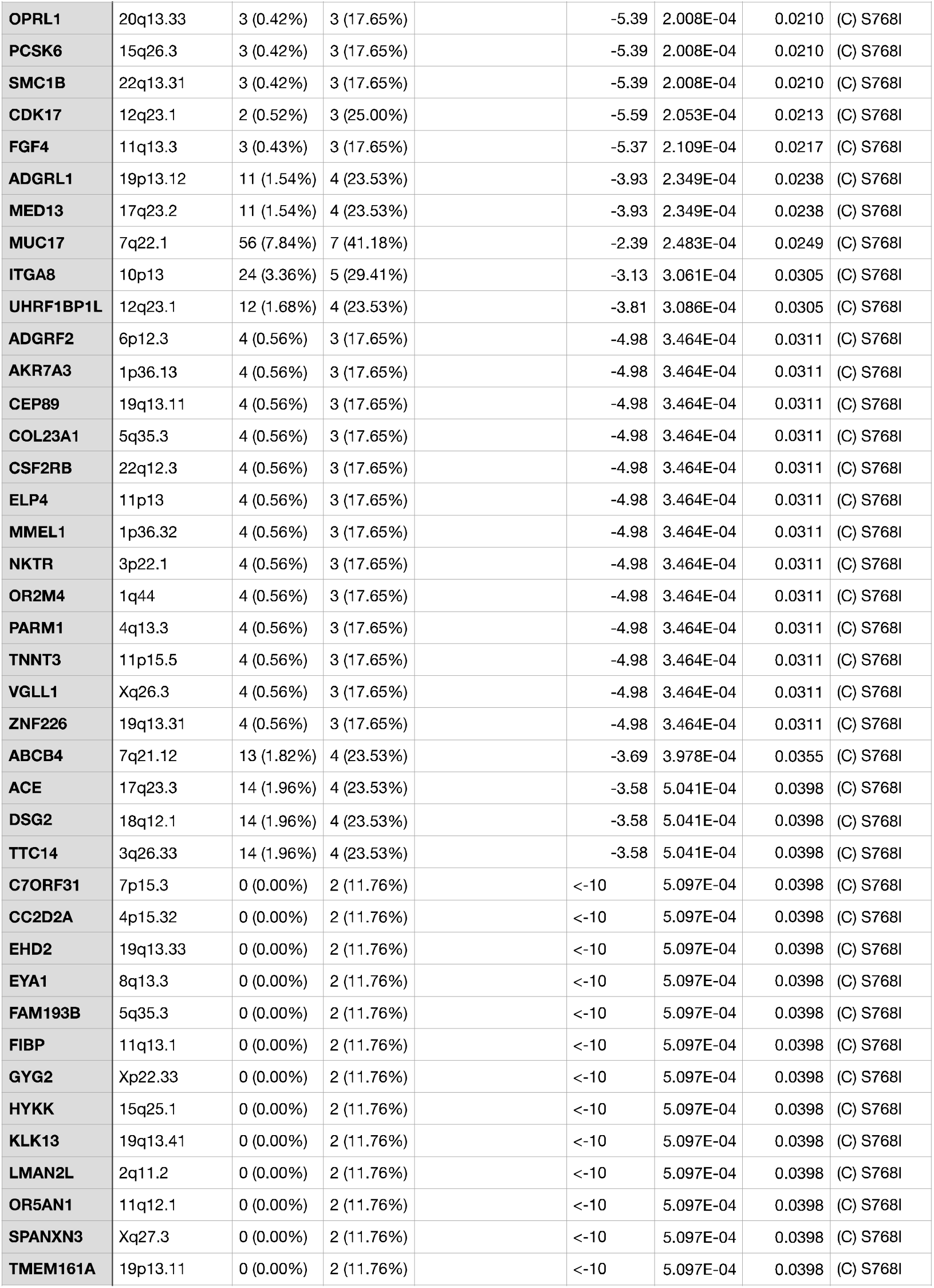
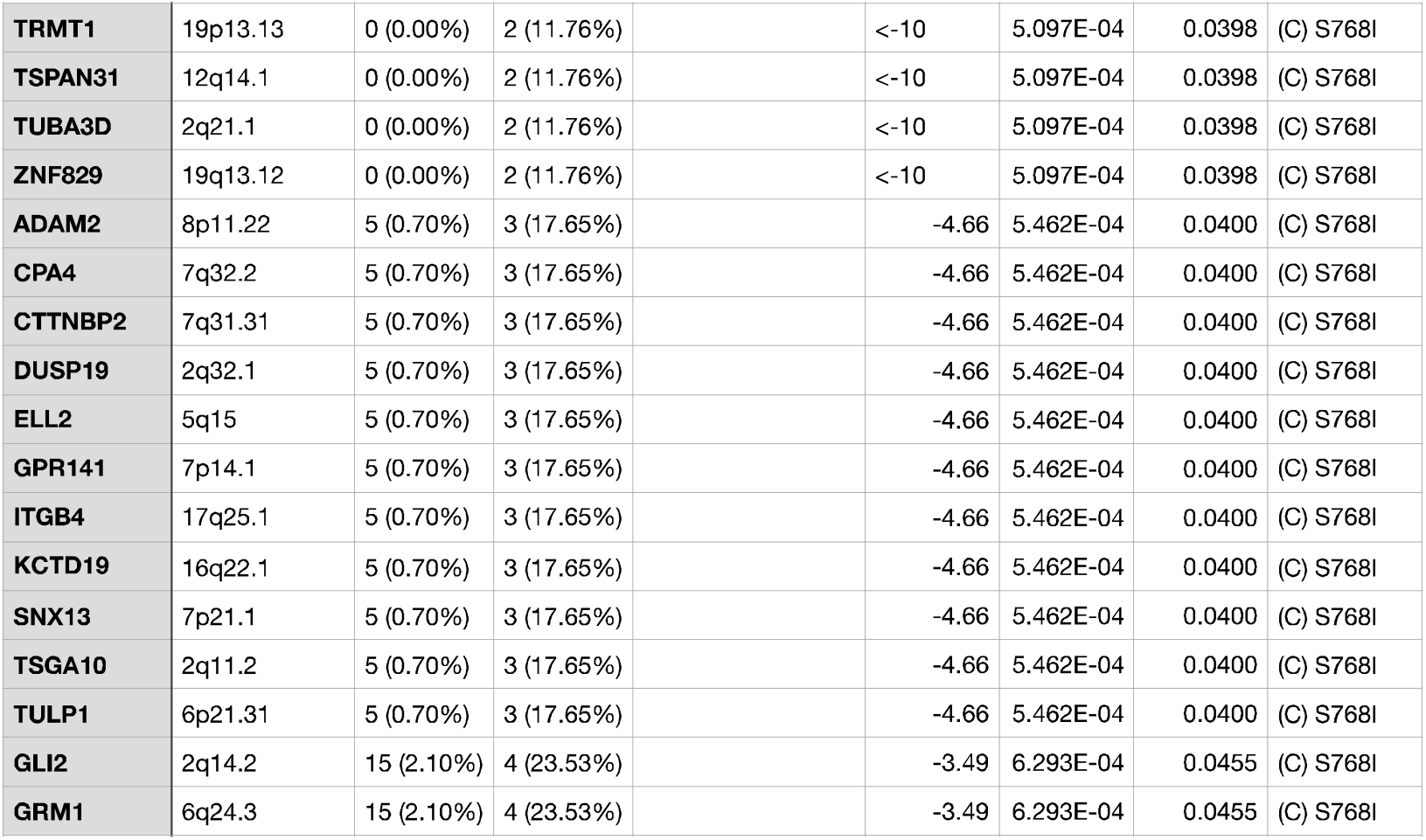

